# Prevalence and risk factors for in-school transmission of SARS-CoV-2 in Massachusetts K-12 public schools, 2020-2021

**DOI:** 10.1101/2021.09.22.21263900

**Authors:** Sandra B. Nelson, Caitlin M. Dugdale, Alyssa Bilinski, Duru Cosar, Nira R. Pollock, Andrea Ciaranello

**Affiliations:** Division of Infectious Diseases, Massachusetts General Hospital, Boston; Medical Practice Evaluation Center, Massachusetts General Hospital, Boston; Department of Laboratory Medicine, Boston Children’s Hospital, Boston; Harvard Medical School, Boston; Department of Health Services, Policy and Practice and Department of Biostatistics, Brown School of Public Health, Providence, Rhode Island

**Author notes:** **Corresponding Author:** Sandra B. Nelson, MD, Division of Infectious Diseases, Massachusetts General Hospital, Cox building, 5^th^ floor, Boston, MA 02114.

## Abstract

**Introduction:** The SARS-CoV-2 secondary attack rate (SAR) in schools is low when mitigation measures are adopted, Data on the relative impact of such strategies are limited. We evaluated the SARS-CoV-2 SAR in Massachusetts schools during 2020-21 and factors associated with transmission risk.

**Methods:** In a convenience sample of 25 Massachusetts public K-12 school districts, de-identified information about SARS-CoV-2 cases and their school-based contacts was reported using a standardized contact-tracing tool. Index cases were included if they were in school while infectious. SAR was defined as the proportion of in-school contacts acquiring SARS-CoV-2 infection and designated as possible or probable in-school transmission by school-based teams. We compared exposure-specific SAR using unadjusted risk ratios (RR) with 95% confidence intervals (CI); p-values were calculated using Fishers exact tests.

**Results:** Eight districts (70 schools with >33,000 enrolled students) participated. There were 435 index cases and 1,771 school-based contacts (Table 1). Most contacts (1327/1771 [75%]) underwent SARS-CoV-2 testing and 39/1327 (2.9%) contacts tested positive. Of 39 positive contacts, 10 (25.6%) had clear out-of-school exposures and were deemed not in-school transmissions, so were excluded from further calculations. Twenty-nine (74.4%) contacts were deemed possible or probable in-school transmissions, resulting in an in-school SAR of 2.2%. Of the 29 in-school transmissions, 6 (20.7%) were staff-to-staff, 7 (24.1%) were staff-to-student, 3 (10.3%) were student-to-staff, and 13 (44.8%) were student-to-student; 6 (20.7%) occurred from index cases attending work/school while symptomatic. The unadjusted SAR (Table 2) was significantly higher if the index case was a staff member versus a student (RR 2.18, 95% CI 1.06-4.49; p=0.030), if the index case was identified via in-school contact tracing versus via school-based asymptomatic testing (RR 8.44, 95% CI 1.98-36.06; p=0.001), if the exposure occurred at lunch versus elsewhere (RR 5.74, 95% CI 2.11-15.63; p<0.001; all lunch transmissions were staff-to-staff), and if both parties were unmasked versus both masked (RR 6.98, 95% CI 3.09-15.77; p<0.001). For students, SAR did not differ by grade level.

**Conclusions:** Secondary attack rates for SARS-CoV-2 were low in public school settings with comprehensive mitigation measures in place before the emergence of the delta variant; lack of masking and staff-to-staff dining were associated with increased risk.

## Introduction

The SARS-CoV-2 secondary attack rate (SAR) in schools is low when mitigation measures are adopted, including masking, ventilation, handwashing, exclusion of symptomatic individuals, screening testing, and isolation and quarantine for those infected and exposed.^1^ Data on the relative impact of such strategies are limited. We evaluated the SARS-CoV-2 SAR in Massachusetts schools during 2020-21 and factors associated with transmission risk.

## Methods

A convenience sample of 25 Massachusetts public K-12 school districts was invited to participate. De-identified information about SARS-CoV-2 cases and their school-based contacts was reported using a standardized contact-tracing tool. Index cases were included if they were in school while infectious.

SAR was defined as the proportion of in-school contacts acquiring SARS-CoV-2 infection and designated as “possible” or “probable” in-school transmission by school-based teams. We compared exposure-specific SAR using unadjusted risk ratios (RR) with 95% confidence intervals (CI); p-values were calculated using Fisher’s exact tests. Full methodologic details are included in Supplemental Material The study was approved by the Mass General Brigham Institutional Review Board.

## Results

Eight districts (70 schools with >33,000 enrolled students) participated. There were 435 index cases and 1,771 school-based contacts (Table 1). Most contacts (1327/1771 [75%]) underwent SARS-CoV-2 testing and 39/1327 (2.9%) contacts tested positive. Of 39 positive contacts, 10 (25.6%) had clear out-of-school exposures and were deemed not in-school transmissions, so were excluded from further calculations.

**Table 1:** Number of SARS-CoV-2 index cases, possible and probable in-school transmissions, secondary attack rates, and number of contacts per index case in 8 Massachusetts K-12 public school districts, 2020-21.

|  |  |  |  |  | In-school transmissions<sup>1</sup> |  |  | Secondary attack rate<sup>2</sup> |  |  | Number of contacts per index case<sup>3</sup> |  |
| --- | --- | --- | --- | --- | --- | --- | --- | --- | --- | --- | --- | --- |
| District | Student enrollment | Index cases<sup>4</sup> | In-school contacts<sup>5</sup> | Contacts tested N (%) | Possible | Probable | Total | Ascertained | Lower bound (assume untested are negative) | Upper bound (assume untested are positive) | Mean (SD) | Median (IQR) |
| 1 | 1000-5000 | 27 | 27 | 25 (93%) | 2 | 0 | 2 | 8.0% | 7.4% | 14.8% | 1.0 (2.0) | 0 (0, 1) |
| 2 | 1000-5000 | 5 | 100 | 99 (99%) | 1 | 0 | 1 | 1.0% | 1.0% | 2.0% | -- | -- |
| 3 | > 5000 | 71 | 416 | 233 (56%) | 0 | 2 | 2 | 0.9% | 0.5% | 44.5% | 5.9 (5.5) | 4 (2, 9) |
| 4 | 1000-5000 | 20<sup>6</sup> | 89 | 89 (100%) | 1 | 0 | 1 | 1.1% | 1.1% | 1.1% | 4.5 (6.0) | 3 (1, 5) |
| 5 | > 5000 | 10 | 73 | 67 (92%) | 5 | 3 | 8 | 11.9% | 11.0% | 19.2% | -- | -- |
| 6 | > 5000 | 165 | 358 | 223 (62%) | 3 | 6 | 9 | 4.0% | 2.5% | 40.2% | 2.4 (5.8) | 1 (0, 2) |
| 7 | < 1000 | 20 | 9 | 9 (100%) | 0 | 0 | 0 | 0.0% | 0.0% | 0.0% | 0.5 (0.9) | 0 (0, 1) |
| 8 | 1000-5000 | 117 | 699 | 582 (83%) | 3 | 3 | 6 | 1.0% | 0.9% | 17.6% | 5.6 (7.1) | 2 (0, 10) |
| **TOTALS** |  | **435** | **1771** | **1327 (75%)** | **15** | **14** | **29** | **2.2%** | **1.6%** | **26.7%** | **3.8 (6.1)** | **1 (0, 5)** |

<sup>1</sup> As adjudicated by the districts reporting the data.
<sup>2</sup> Secondary attack rate was calculated in three ways - as ascertained by testing (the primary analysis: number of contacts testing positive divided by number of contacts tested), as a lower bound (assuming all untested contacts were truly uninfected), and as an upper bound (assuming all untested contacts were truly infected).
<sup>3</sup> Mean and median contacts per index case were only calculated for those districts that provided information on all cases who were present in school during the infectious window, including cases with zero contacts. Districts 2 and 5 provided data only for index cases with at least one contact, and thus were excluded from the analyses of mean and median contacts per case.
<sup>4</sup> Index cases were only included if they were present in school during the infectious window, 48 hours prior to symptom onset (or positive test if asymptomatic) until the time they were isolated. The number of index cases include cases in which there were zero in-school contacts, but the case was present in school during the infectious window.
<sup>5</sup> Contacts were defined as those individuals within 6 feet for at least 15 minutes over 24 hours during the infectious window, unless the contact was fully vaccinated. In April 2021, those close contacts within classrooms and on school busses were excluded from quarantine requirements if both the case and contact were masked, unless they were closer than 3 feet together for at least 15 minutes over 24 hours during the infectious window.
<sup>6</sup> This district provided data only on cases who were identified through school-based asymptomatic testing protocols.

Twenty-nine (74.4%) contacts were deemed possible or probable in-school transmissions, resulting in an in-school SAR of 2.2%. Of the 29 in-school transmissions, 6 (20.7%) were staff-to-staff, 7 (24.1%) were staff-to-student, 3 (10.3%) were student-to-staff, and 13 (44.8%) were student-to-student; 6 (20.7%) occurred from index cases attending work/school while symptomatic.

The unadjusted SAR (Table 2) was significantly higher if the index case was a staff member versus a student (RR 2.18, 95% CI 1.06-4.49; p=0.030), if the index case was identified via in-school contact tracing versus via school-based asymptomatic testing (RR 8.44, 95% CI 1.98-36.06; p=0.001), if the exposure occurred at lunch versus elsewhere (RR 5.74, 95% CI 2.11-15.63; p<0.001; all lunch transmissions were staff-to-staff), and if both parties were unmasked versus both masked (RR 6.98, 95% CI 3.09-15.77; p<0.001). For students, SAR did not differ by grade level.

**Table 2:** Number of index cases and contacts and secondary attack rate by type of exposure: 8 public MA K-12 districts, 2020-21.

| Exposure characteristics | # Index cases | Total # contacts | # Contacts tested | # In-school transmissions | SAR (%)<sup>1</sup> | RR (95% CI) | p-value | Mean contacts/case<sup>2</sup> |
| --- | --- | --- | --- | --- | --- | --- | --- | --- |
| **Contact role** |  |  |  |  |  |  |  |  |
| Student | 216 | 1492 | 1093 | 20 | 1.8 | Ref | ---- | 6.56 |
| Staff | 151 | 278 | 233 | 9 | 3.9 | 2.11 (0.97, 4.58) | 0.054 | 1.79 |
| **Contact grade level** |  |  |  |  |  |  |  |  |
| PK-5 | 119 | 798 | 633 | 12 | 1.9 | Ref | ---- | 6.45 |
| 6-8 | 40 | 205 | 136 | 3 | 2.2 | 1.16 (0.33, 4.07) | 0.812 | 4.47 |
| 9-12 | 36 | 328 | 255 | 4 | 1.6 | 0.83 (0.27, 2.54) | 0.740 | 8.67 |
| **Index role<sup>3</sup>** |  |  |  |  |  |  |  |  |
| Student | 286 | 1276 | 967 | 16 | 1.7 | Ref | ---- | 4.06 |
| Staff | 149 | 495 | 360 | 13 | 3.6 | 2.18 (1.06, 4.49) | 0.030 | 3.30 |
| **Index grade level<sup>3</sup>** |  |  |  |  |  |  |  |  |
| PK-5 | 107 | 636 | 518 | 8 | 1.5 | Ref | ---- | 5.68 |
| 6-8 | 70 | 224 | 145 | 3 | 2.1 | 1.34 (0.36, 4.99) | 0.662 | 2.76 |
| 9-12 | 104 | 414 | 295 | 7 | 2.4 | 1.54 (0.56, 4.19) | 0.399 | 3.64 |
| **Mechanism for identifying the index case<sup>3</sup>** |  |  |  |  |  |  |  |  |
| School-based asymptomatic testing protocol | 100 | 384 | 325 | 7 | 2.2 | Ref | ---- | 3.64 |
| Tested because close contact IN school | 6 | 13 | 11 | 2 | 18.2 | 8.44 (1.98, 36.06) | 0.001 | 2.00 |
| Tested because close contact OUT of school | 128 | 588 | 413 | 5 | 1.2 | 0.56 (0.18, 1.75) | 0.315 | 4.61 |
| Tested before or after travel | 5 | 20 | 9 | 0 | 0.0 | n/a | 0.656 | 4.00 |
| Tested for symptoms | 194 | 766 | 569 | 15 | 2.6 | 1.22 (0.50, 2.97) | 0.654 | 3.42 |
| **Mechanism for identifying the index case** |  |  |  |  |  |  |  |  |
| School-based asymptomatic testing protocol | 100 | 384 | 325 | 7 | 2.2 | Ref | ---- | 3.64 |
| Other | 333 | 1387 | 1002 | 22 | 2.2 | 1.02 (0.44, 2.36) | 0.964 | 3.88 |
| **Location of contact: Classroom** |  |  |  |  |  |  |  |  |
| No known classroom exposure | 114 | 480 | 368 | 12 | 3.3 | Ref | ---- | 3.94 |
| Classroom exposure | 187 | 1291 | 959 | 17 | 1.8 | 0.54 (0.26, 1.13) | 0.097 | 6.71 |
| **Classroom exposures: index role** |  |  |  |  |  |  |  |  |
| Classroom exposure with staff index | 54 | 358 | 245 | 7 | 2.9 | Ref | ---- | 6.86 |
| Classroom exposure with student index | 133 | 933 | 714 | 10 | 1.4 | 0.49 (0.19, 1.27) | 0.136 | 6.65 |

**Table 2: Number of index cases and contacts and secondary attack rate by type of exposure: 8 public MA K-12 districts, 2020-21, continued.**
| Exposure characteristics | # Index cases | Total # contacts | # Contacts tested | In-school transmissions | SAR (%)<sup>1</sup> | RR (95% CI) | p-value | Mean contacts/case<sup>2</sup> |
| --- | --- | --- | --- | --- | --- | --- | --- | --- |
| Location of contact: Lunch |  |  |  |  |  |  |  |  |
| No known lunch exposure | 258 | 1728 | 1291 | 25 | 1.9 | Ref | ---- | 6.35 |
| Lunch exposure | 18 | 43 | 36 | 4 | 11.1 | 5.74 (2.11, 15.63) | <0.001 | 2.14 |
| Lunch exposure: index role |  |  |  |  |  |  |  |  |
| Lunch exposure with staff index | 15 | 29 | 27 | 4 | 14.8 | Ref | ---- | 1.45 |
| Lunch exposure with student index | 3 | 14 | 9 | 0 | 0.0 | n/a | 0.221 | 4.67 |
| Location of contact: bus |  |  |  |  |  |  |  |  |
| No known bus exposure | 255 | 1716 | 1289 | 29 | 2.2 | Ref | ---- | 6.45 |
| Bus exposure | 24 | 55 | 38 | 0 | 0.0 | n/a | 0.350 | 2.23 |
| Location of contact: school sports |  |  |  |  |  |  |  |  |
| No known school sports exposure | 254 | 1532 | 1152 | 24 | 2.1 | Ref | ---- | 5.84 |
| School sports exposure | 19 | 239 | 175 | 5 | 2.9 | 1.37 (0.53, 3.55) | 0.514 | 11.59 |
| Mask use between case and contact<sup>4</sup> |  |  |  |  |  |  |  |  |
| Both masked | 250 | 1687 | 1256 | 21 | 1.7 | Ref | ---- | 6.41 |
| Both unmasked | 28 | 67 | 60 | 7 | 11.7 | 6.98 (3.09, 15.77) | <0.001 | 2.25 |

SAR: secondary attack rate; RR: (unadjusted) relative risk, CI: confidence interval.
<sup>1</sup> SAR was calculated based on the number of contacts who were known to have undergone SARS-CoV-2 testing.
<sup>2</sup> Mean contacts/case was calculated based on the total number of reported contacts, irrespective of whether or not those contacts underwent testing, but excluding data from two districts that only reported cases if they had at least one contact.
<sup>3</sup> The number of index cases and mean contacts/case for these analyses include cases who were not known to have any associated contacts in-school. The remainder of the analyses only include number of index cases that had at least one contact, as they examine contact-related exposure characteristics.
<sup>4</sup> There were only 3 contacts who were exposed in settings in which either the case or the contact was masked, but not both individuals, so these data are not shown.

## Discussion

Based on detailed school-based contact tracing, the SARS-CoV-2 SAR among school-based contacts in Massachusetts was low. While other studies have demonstrated the benefit of policies requiring masks in K-12 schools,^2,3^ detailed contextual information about in-school exposures remains necessary to inform policy. This study provides in-depth analysis of transmission context, extending existing literature by highlighting the potential benefit of masking in school settings, the risk of lunchtime adult-to-adult transmission, and the similarity of SARs across ages/grade levels.

Our study has important limitations. Although contact-tracing information was recorded in real time, districts may have inferred missing data inaccurately, with potential for recall bias. These data are derived from the 2020-21 school year, before the emergence of the more-transmissible delta variant. We lacked sufficient data to assess other potentially confounding factors, such as duration of exposure, classroom distancing, ventilation, and classroom density. Notably, all reported classroom exposures were masked, so these results do not directly inform the impact of masking within classrooms.

The generalizability of these data for the 2021-22 year is uncertain. Most schools are now resuming full in-person learning with greater classroom density, reduced distancing, and variable approaches to masking. While community and student/staff vaccination should reduce the number of people entering school buildings with SARS-CoV-2 infection, this impact may be offset by reduced mitigation measures and more transmissible variants.^4^ Ongoing surveillance of in-school SARS-CoV-2 transmission is critical to inform decisions about school-based mitigation measures as the pandemic evolves.

## Data Availability

Data are available from the authors upon appropriate request.

## Acknowledgements

We are indebted to the efforts of the school staff in the districts, who worked tirelessly throughout the pandemic to enable safe school reopening, and who shared their experiences with us through this study. These include Nicole Altieri, Amy Bantham, Lynn Clark, Mary Ellen Duggan, Noelle Freeman, Tricia Laham, Joyce O’Neil, Chassea Robinson, Michelle Shuckel Fronk, Jill Seaman-Chandler, Patricia Smith, Jenny Tam, and Ruth Wagner.

## Supplemental Methods

We created a standardized spreadsheet for reporting de-identified information about COVID-19 cases in schools and their school-based contacts. Index cases were only included if they were in school during the infectious window, defined as 48 hours before symptom onset or before a positive test was collected until the time of isolation. Requested information about the index cases included their role in school (i.e., student/staff and grade level/staff role), the means of case identification (e.g., regularly scheduled asymptomatic testing, symptomatic testing, testing after exposure in school, or testing after exposure outside of school), their duration of time spent in-school while infectious, and their number of in-school close contacts. Contacts were defined by Massachusetts Department of Public Health standards as those individuals within 6 feet of an index case for at least 15 minutes (cumulative) over 24 hours during the infectious window, unless the contact was fully vaccinated.^1^ In April of 2021, close contacts within classrooms and on school buses were excluded from quarantine requirements if both the case and contact were masked, unless closer than 3 feet for at least 15 minutes over 24 hours during the infectious window.^2^ Information about the contacts included their role in school (i.e., student/staff and grade level/staff role), the location of exposure (e.g., classroom, lunch/snack, recess, physical education (PE), bus, school sports, other school-sponsored extracurricular), the presence masking during the exposure, whether or not they were tested, and the results of testing. Additional qualitative information about the circumstances of possible or probable school-based transmission was included. School-based nursing and/or contact tracing teams designated contacts as “not in school transmission” if a clear alternative exposure was present and felt to be more likely than the in-school exposure (i.e., a household contact with exposure timing more convincing for likely source of infection).

Descriptive analyses were performed to calculate the total number of cases, contacts, and possible or probable in-school transmission events for each district and in each category of index case and exposure type (student/staff, grade level, exposure setting, masking, etc.), in addition to the mean/median number of contacts per case and the proportion of contacts who underwent SARS-CoV-2 testing within 14 days of their exposure. We calculated secondary attack rates, defined as the proportion of contacts acquiring SARS-CoV-2 infection and not considered “not in school transmission,” in three ways: 1) as ascertained by testing (the primary analysis: number of contacts testing positive divided by number of contacts tested), 2) a lower bound (assuming all untested contacts were truly uninfected), and 3) an upper bound (assuming all untested contacts were truly infected). When data on the proportion of contacts tested were not available for an individual index case, district health staff estimated the proportion tested; these estimations occurred primarily in the setting of cases involving two large high school sports teams from a single district. Descriptive information about possible and probable school-based transmissions was summarized to calculate secondary attack rates by district and by exposure type.

